# Field performance evaluation of the PanBio rapid SARS-CoV-2 antigen assay in an epidemic driven by 501Y.v2 (lineage B.1.351) in the Eastern Cape, South Africa

**DOI:** 10.1101/2021.02.03.21251057

**Authors:** Oluwakemi Laguda Akingba, Kaitlin Sprong, Diana Ruth Hardie

## Abstract

**Background:** South Africa was the African country most severely affected by the SARS-CoV-2 pandemic during 2020, experiencing 2 waves of infection. During the first wave, diagnostics were largely based on reverse transcription-linked PCR (RT-PCR). The Abbott PanBio antigen test was deployed during the 2nd wave which was driven by emergence of the 501Y.v2 variant. At the time of evaluation in mid-November 2020, 501Y.v2 was the dominant circulating virus in Nelson Mandela Bay, in the Eastern Cape Province.

**Methods:** A prospective diagnostic evaluation study was undertaken, during a period of high community transmission, to evaluate the field performance of the PanBio antigen RTD. Testing was conducted at mobile community testing centres on 677 ambulant patients seeking SARS-CoV-2 testing. RT-PCR was performed on the original naso-pharyngeal antigen swabs to evaluate test performance.

**Results:** Of 146 RT-PCR positive individuals, 101 were RTD positive in the clinic. The antigen RTD had an overall sensitivity of 69.2% (95%CI 61.4, 75.8) and specificity of 99.0% (95%CI 98.8, 99.3) in this clinical context. Sensitivity was strongly dependent on the amount of virus in clinical samples, as reflected by the PCR cycle threshold (CT) value, with 100% detection in samples where the CT was <20, 96% with CT between 20-25, 89% with CT between 26-30 and 64% when CT was 31-35.

**Conclusions:** The assay reliably detected 501Y.v2 infections in ambulatory ill patients. Assay sensitivity was >90% in patients with high viral loads who are expected to be most infectious. Negative and positive predictive values were also >90%.

## Background

During 2020 South Africa was the African country most severely affected by the SARS-CoV-2 pandemic with more than 1 380 000 laboratory confirmed cases and 83 918 excess deaths[1]. During this period, the country experienced 2 waves of infection[2]. Provision of an effective diagnostic service proved to be challenging. RT-PCR is the gold standard assay for SARS-CoV-2 diagnosis, however, in the context of high disease prevalence, laboratory systems may easily become overwhelmed. Rapid diagnostics such as antigen tests that can be performed at point of care provide a welcome solution. Their main drawback is lower sensitivity[3]. The WHO advises that assays that meet minimum performance requirements (>80% sensitivity, >97% specificity in the first 7 days of symptoms) can be used in contexts where nucleic acid-based testing is unavailable, or where turn-around times are prolonged[4]. The Abbott PanBio rapid SARS-CoV-2 antigen assay has fulfilled these criteria in evaluations in several studies [5][6]

This assay was deployed during the 2^nd^ wave in South Africa which first became apparent in the Eastern Cape Province in October/November 2020. Increased disease activity was associated with emergence of a new variant, namely 501Y.v2[7]. This variant, now referred to as lineage B.1.351, first detected in October 2020 rapidly became the predominant virus, across the country, due to its higher transmissibility[8]. At the time of evaluation in mid-November 2020, it was the dominant circulating virus, responsible for around 84% of infections in Nelson Mandela Bay, estimate based on genomes submitted to global initiative for sharing all influenza data (GISAID) over this time period [9]. This prospective diagnostic evaluation study was designed to evaluate the field performance of the PanBio assay, but also provides evidence on its performance in individuals infected with 501Y.v2. Another novel aspect to this study is that RT PCR was performed on the same swab used for antigen testing, which obviated the need to collect further samples from patients and provided a more direct comparison with the antigen result.

## Methods and results

Prospective diagnostic evaluation study in Nelson Mandela Bay municipality, Eastern Cape South Africa during a period of high disease prevalence, using nasopharyngeal swabs to determine the accuracy of Abbott PanBio COVID-19 antigen RTD.

### Verifying that used antigen swabs were suitable for PCR

44 paired swabs were collected from symptomatic patients, one nylon tip, standard issue swab for PCR and the flocked antigen swab from the PanBio test kit. SARSCoV-2 PCR was done on used antigen and matched nylon swabs.

The used antigen swab was prepared for PCR as follows: 1 mL saline was added to the swab container using a filter tip. The sample was vortexed and allowed to stand 2 minutes. The bottom cap was opened and fluid bled into a sterile vial. The matching PCR swab was snipped into a vial containing 1.5 ml normal saline and vortexed.

Paired samples were extracted on the EasyMag (bioMerieux) platform. RT-PCR was done using the Seegene nCoV assay with amplification on BioRad CFX realTime PCR machine. For PCR positive swabs, mean CT values of the 3 assay targets were compared.

Of the 44 paired samples, 13 were antigen positive. 26 were concordantly PCR negative, one sample was PCR positive on the antigen swab, but negative on the regular swab and 17 samples were concordantly PCR positive. When comparing mean CT values of the paired swabs, the antigen swab had values of 2 CTs lower than the swab collected for PCR. Wilcoxon signed-rank test p0.0073. (figure 1)

**Figure 1:**
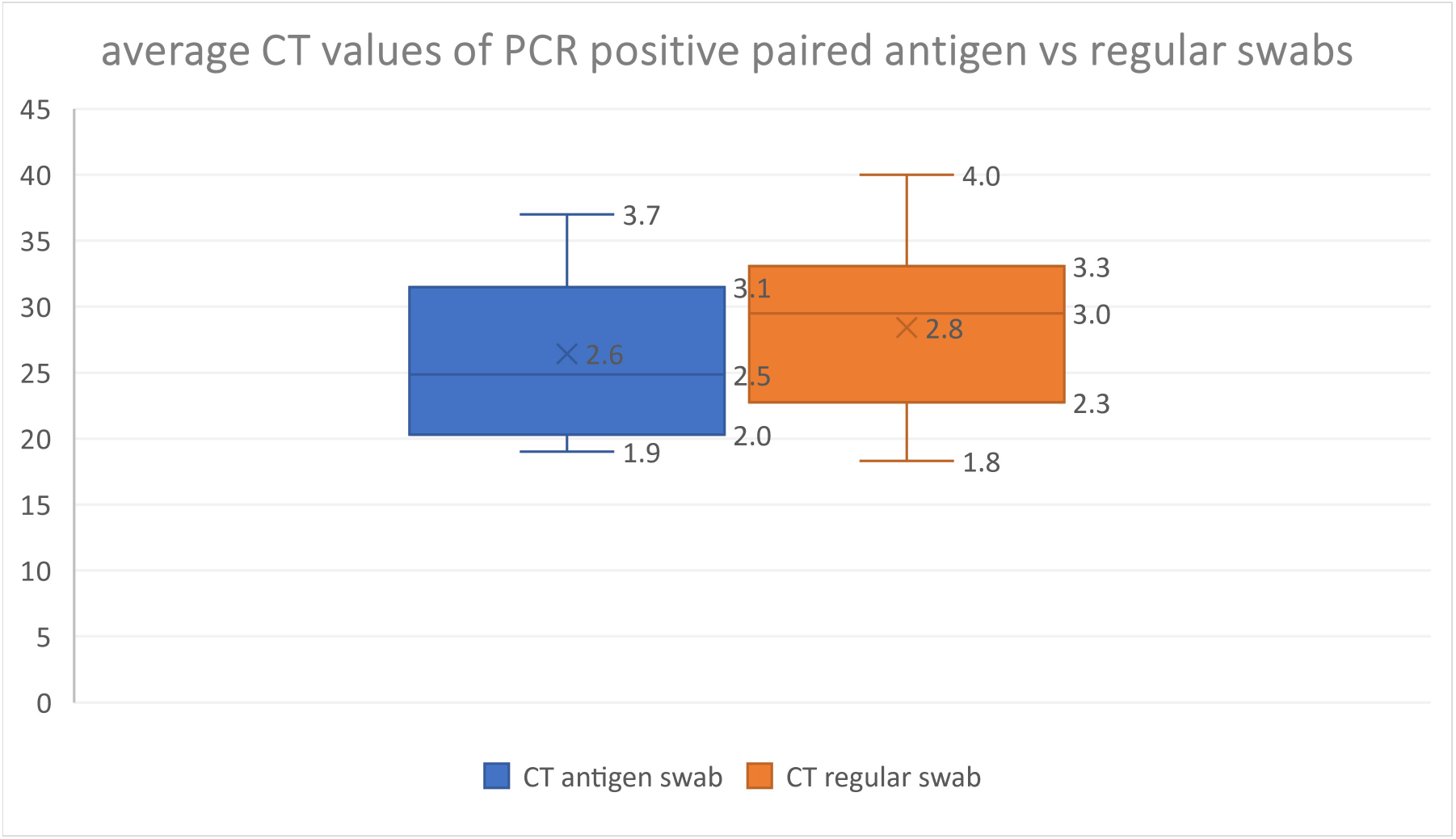
Average CT values of PCR positive paired antigen vs regular swabs are compared: The range of CT values from PCR of the antigen swab were on average 2 CTs lower than for those from the standard swabs. Wilcoxon signed-rank test p0.0073.

### Study protocol

Between 17 and 20 November 2020 mobile clinics ran community testing campaigns at 6 sites in Nelson Mandela Bay. Symptomatic patients were invited to undergo antigen testing. Nasopharyngeal (NP) swabs were tested using the PanBio SARS-CoV-2 RTD. Results were communicated to patients immediately. The used swabs were sent for PCR. RT-PCR: Seegene nCoV assay (BioRad CFX platform) was performed on the same swab used for antigen testing. (as described)

A total of 677 patients from 6 mobile clinics were tested by both antigen and PCR. Patients were ambulant and seeking COVID testing. They ranged in age range from 3-85 years; 59% were female.

Of these, 101 (14.9%) were antigen positive in the clinic. With PCR, 146 samples (21.4%) were reported as positive, 19 (2.8%) as inconclusive (single target positive, CT>38) and 509 (75.2%) were negative for both tests. Inconclusive samples were excluded from analysis as their significance was unresolved.

### Antigen test performance

Using PCR as the reference standard, the antigen test had an overall sensitivity (positive percent agreement) of 69.17% (95%CI 61.44, 75.80) and specificity (negative percent agreement) of 99.02% (95%CI 98.78, 99.26) in this clinical context.

Sensitivity was strongly dependent on the quantity of virus in clinical samples, as reflected by the CT value, with 100% detection by the antigen test in samples where the CT was <20, 95.5% with CT between 20-25, 89.3% with CT between 26-30 and 64,3% when CT was 31-35. The CT values of antigen positive and negative samples are shown (Figures 2 a, b)

**Figure 2.**
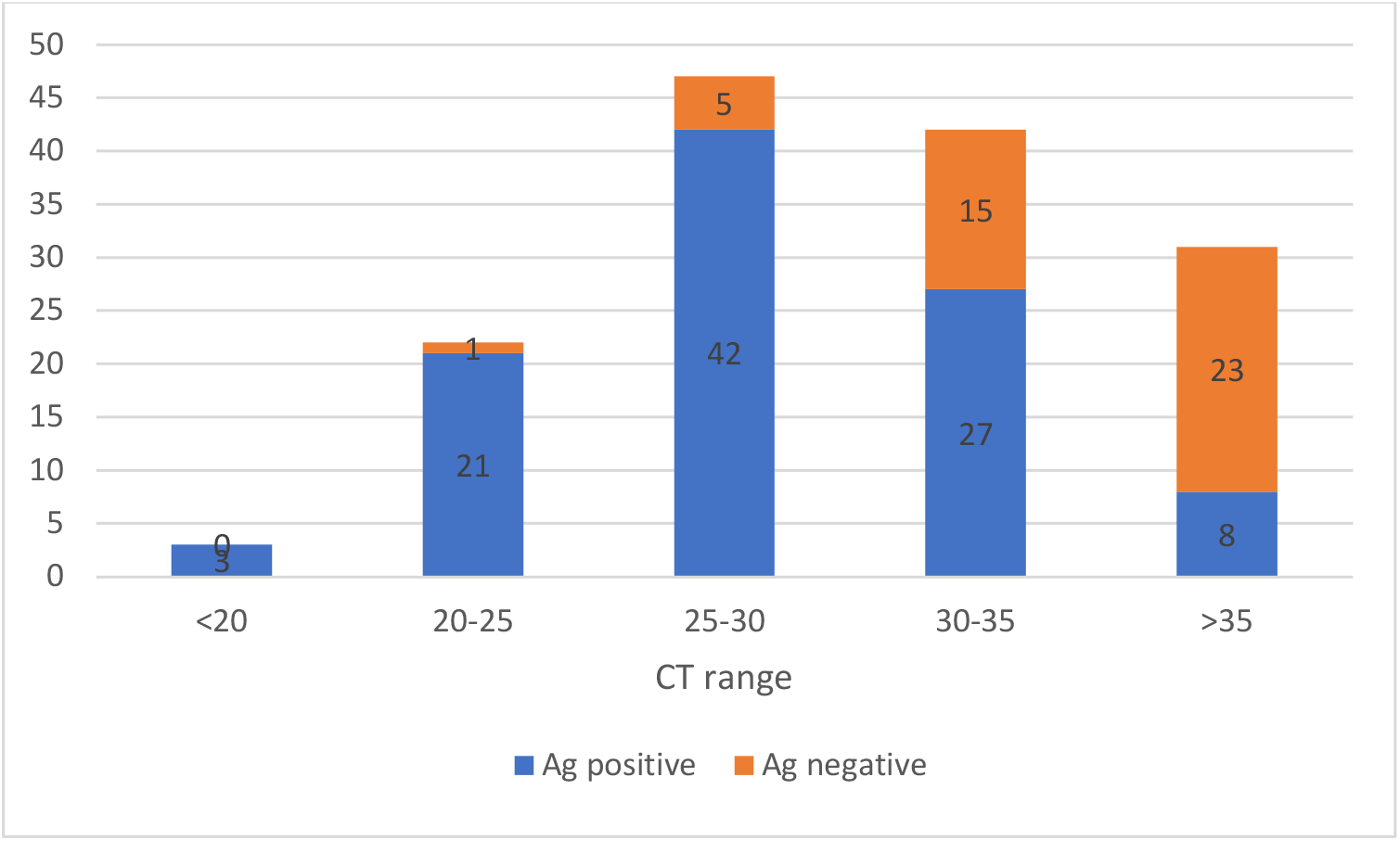
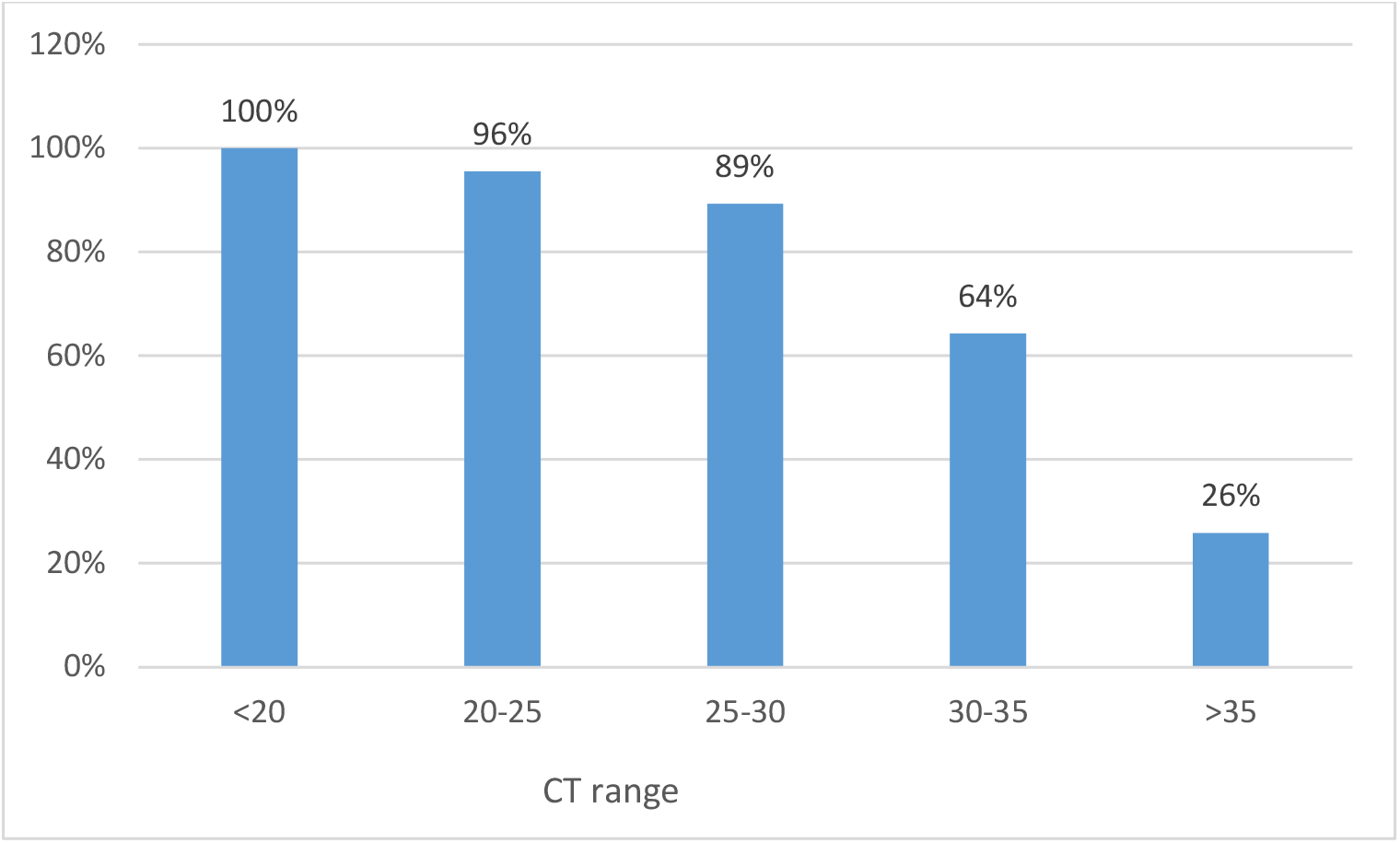
(a) Compares the number of antigen positive and negative samples according to CT category values obtained in PCR, (b) Compares the percentage of antigen positive samples according to virus levels, as reflected by the mean CT value.

The antigen assay was positive in 3 PCR negative patients. Given the prevalence of infection of 21% (as determined by PCR) the predictive value of a negative test was 91.9% and that of a positive test was 97.12%.

### Characteristics of PCR positive samples

In the 146 PCR positive patients, CT values ranged from 17.4 to 41.3, median 30.1. As expected, the median CT and interquartile range (IQR) was lower in antigen positive samples at 28.7 (IQR 25.3-31.3). In comparison, the median and IQR of CT of antigen negatives was 35.8 (IQR 32.7-37.1). (figure 3)

**Figure 3:**
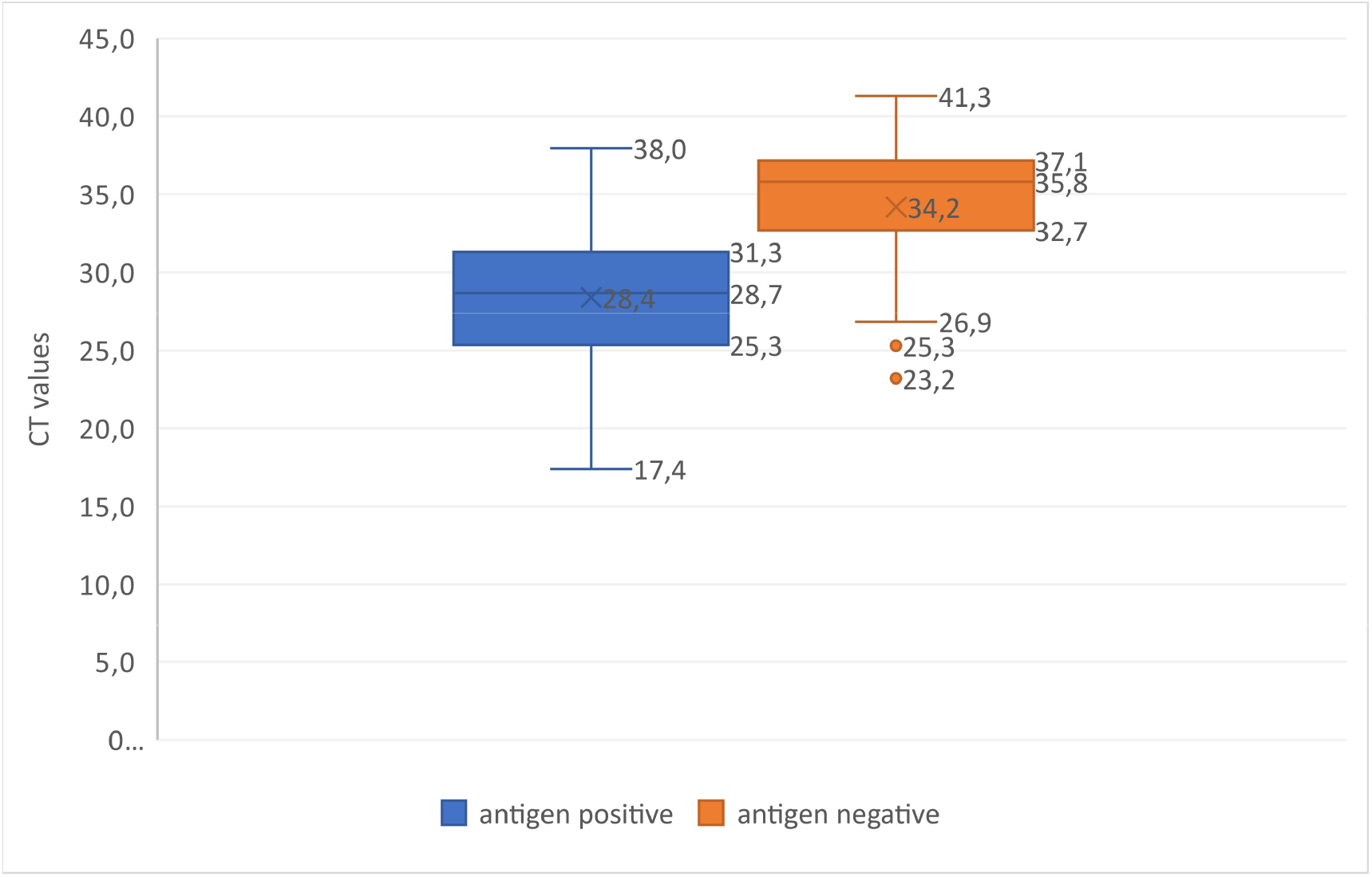
Compares the range of CT values obtained on antigen positive and negative samples by reference PCR. Median and IQR values are given.

## Discussion

This study took place during a period of high community transmission associated with emergence of the 501Y.v2 variant in the Eastern Cape. PCR was performed directly on the antigen swab after testing which enabled a direct comparison to be made between antigen reactivity and PCR on the same sample. Overall, the PanBio antigen test had a sensitivity of 69.17% and specificity of 99.0%. The sensitivity is below the 80% WHO benchmark[4]. However, context is key. Testing was performed on unselected symptomatic individuals who requested testing, irrespective of symptom duration. This probably accounts for the fact that 50% of PCR positive patients had CT values > 30, indicating that on average sampling may have occurred later during infection than recommended for maximum performance. Nonetheless performance was good in patients with CTs <30. In this range, sensitivity was 91.3%. This concordance (using distinctly different technologies) suggests that the 501Y.v2 was reliably detected by the RTD and at an expected level of frequency when compared with PCR.

Assay specificity was similarly good at 99% and the predictive value of a positive test was 95%. This fulfils the WHO benchmark specificity requirements for deployment of this assay[4].

2 factors could compromise detection of the 501Y.v2 variant; namely amino acid changes in the region of the nucleocapsid protein targeted by kit antibodies, or reduced virus shedding in respiratory tract samples. The higher infectivity of the variant makes the latter explanation unlikely and preliminary evidence does not support it. 501Y.v2 has a single amino acid change in the linker region of the nucleocapsid protein, namely N205I [9]. As this amino acid is located in an unstructured region[10], it should not affect binding of kit antibodies.

The main limitation was that it was not feasible to confirm 501Y.v2 infection in positive cases. This was inferred from the fact that resurgence in this district was overwhelmingly due to 501Y.v2, based on contemporaneous genomes submitted to GISAID [9].

## Conclusion

The assay reliably detected 501Y.v2 virus infection in ambulatory ill patients in this high prevalence community setting. Sensitivity was >90% in patients with high viral loads who are expected to be most infectious. To optimise the use of antigen RDTs in different and changing circumstances, clinical predictors and the epidemiological context should be considered when deciding how to deploy these assays.

### Ethics

Ethical approval was obtained from the University of Cape Town Human Research Ethics Committee (UCT HREC 862.2020).

### Informed consent

Patients were willing participants and gave verbal consent to diagnostic testing. Data was anonymised and delinked.

## Data Availability

data is available on request from corresponding author

## Author contributions

Study concept and design: DH, OL

Conduct of study, sample testing: OL, KS

Manuscript write up: DH, OL, KS

All authors approved the final manuscript

## Declaration of competing interest

The authors report no competing interests.

## Funding

No funding was given for this study.

## Abbreviations

COVID-19: Coronavirus disease 2019;
CT: cycle threshold;
SARS-CoV-2: severe acute respiratory syndrome coronavirus 2;
RTD: rapid test device;
PCR: polymerase chain reaction;
RT-PCR: reverse transcription polymerase chain reaction;
WHO: world health organisation;
501Y.v2: 501Y variant 2;
IQR: interquartile range;
NP: nasopharyngeal;
GISAID: global initiative for sharing all influenza data;

## Highlights

- **PanBio SARS-CoV-2 RTD performs well in a field evaluation with high disease prevalence**
- **Good concordance between RT-PCR and RTD in patients with 501Y**.**v2 variant infection**.
- **RT-PCR on the antigen swab enables a more direct comparison of methods**.

## Notes

### Competing Interest Statement

The authors have declared no competing interest.

### Funding Statement

no funding was provided for this evaluation. it was performed as a method verification exercise by National health laboratory service in South Africa

### Author Declarations

University of Cape Town human research ethics committee UCT HREC 862.2020

